# Molecular beacons allow specific RT-LAMP detection of B.1.1.7 variant SARS-CoV-2

**DOI:** 10.1101/2021.03.25.21254356

**Authors:** Scott Sherrill-Mix, Gregory D. Van Duyne, Frederic D. Bushman

## Abstract

Over the course of the COVID-19 pandemic, several SARS-CoV-2 genetic variants of concern have appeared and spread throughout the world. Detection and identification of these variants is important to understanding and controlling their rapid spread. Current detection methods for a particularly concerning variant, B.1.1.7, require expensive qPCR machines and depend on the absence of a signal rather than a positive indicator of variant presence. Here we report an assay using a pair of molecular beacons paired with reverse transcription loop mediated amplification to allow isothermal amplification from saliva to specifically detect B.1.1.7 and other variants which contain a characteristic deletion in the gene encoding the viral spike protein. This assay is specific, affordable and allows multiplexing with other SARS-CoV-2 LAMP primer sets.

## Introduction

SARS-CoV-2 virus was first detected in December of 2020 and rapidly spread throughout the world. To date, the virus has infected over 120 million people and caused over 2.5 million deaths. Over the course of the pandemic, several genetic variants of concern have appeared and rapidly increased in frequency. The effects of these mutations remain unclear but there is concern that these variants may alter virus phenotype, affect detection and escape immune responses ^1–10^.

Several variants of concern, including the B.1.1.7 variant first identified in the UK, have a characteristic six nucleotide deletion in the spike gene (S1Δ69-70) that appears likely to be an effective target for detection by nucleic acid testing. Detection of these variants currently requires observation of the dropout of fluorescence from a spike-targeted primer-probe set during qPCR ^1^. However, screening using the absence of a signal is prone to falsely label samples near the limit of detection or assay failures as variants and performing qPCR requires costly RT-qPCR machines. Here, we design a molecular beacon spanning this S1Δ69-70 deletion that allows characterization of virus using simple isothermal reverse transcription-loop mediated amplification (RT-LAMP).

## Materials and Methods

To initiate LAMP reactions, primers are designed such that the ends of the synthesized DNA will form two dumbbell-like loops of single stranded DNA. The single stranded sequence in these loops is dependent on the template DNA and can be completely independent of the primers input to the reaction. Thus this sequence is resistant to creation by artifactual off-target amplification and available for annealing, making it a favorable target for molecular beacons. We used the Primer Explorer software to design a LAMP primer set (S6970) spanning the S1Δ69-70 deletion such that the deletion falls within the backward loop region of the amplification (Table 1) and then designed a molecular beacon (S6970 B117 MB) to both land inside the loop and span the deletion. We also added locked nucleic acids to increase specificity and strengthen hybridization to allow real time detection at the temperatures used in LAMP. In addition, we synthesized an additional molecular beacon (S6970 WT MB) targeting the undeleted S1 69-70 sequence to allow parallel detection of strains containing the preexisting spike sequence (Figure 1A).

**Table 1:**
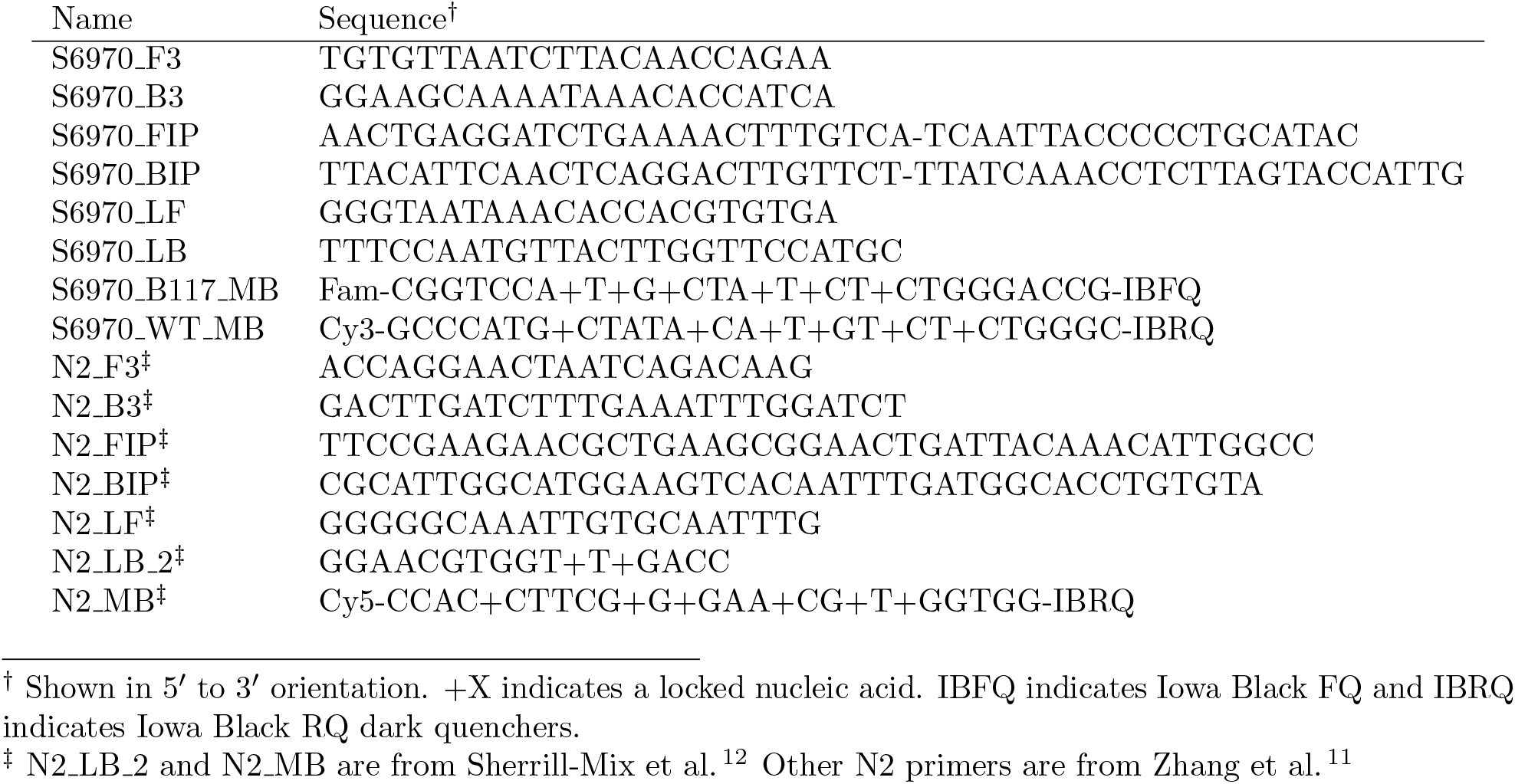
Oligonucleotides used in this study.

**Figure 1.**
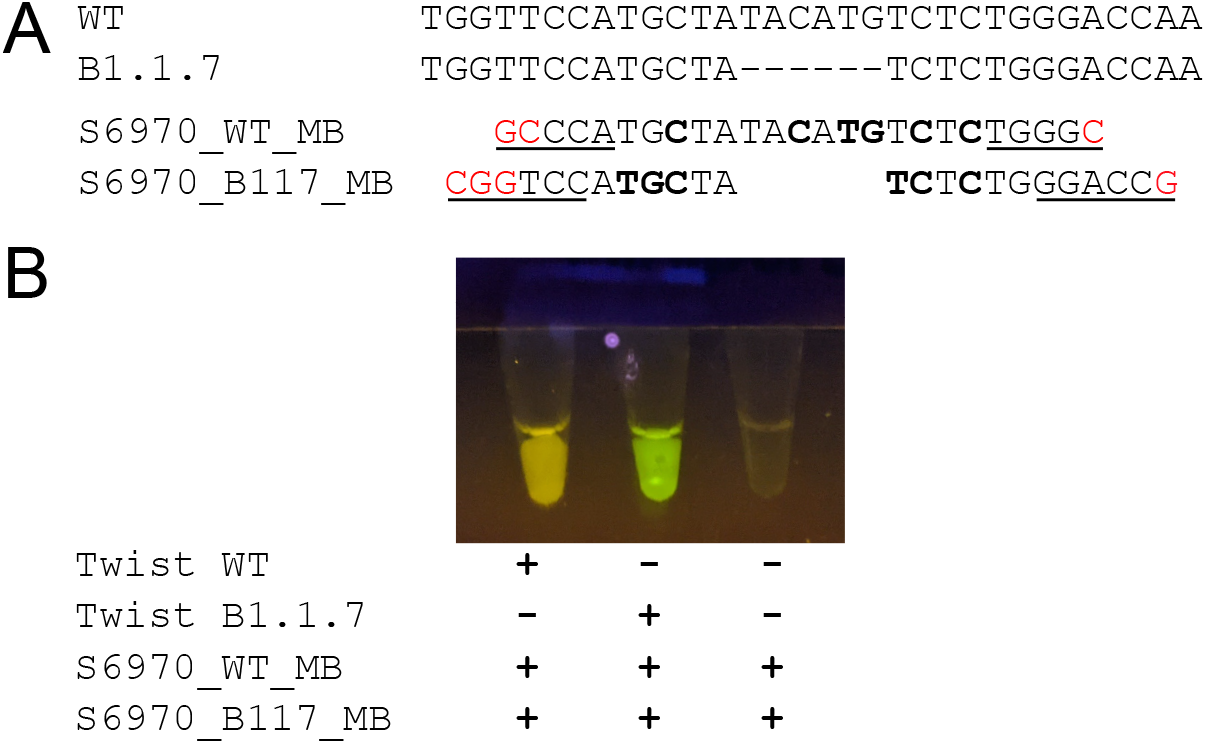
Development of molecular beacons to discriminate SARS-CoV-2 variants with a S1Δ69-70 mutation. A) A comparison of preexisting genomic S1 sequence (WT) and variant B.1.1.7 genomic sequence (B.1.1.7) and molecular beacons targeting these sequences (S6970 WT MB and S6970 B117 MB, respectively). Dashes indicate the S1Δ69-70 deletion, bold letters indicate locked nucleic acids, underlined letters indicate bases paired in the stem of the beacon and red letters indicate extra nucleotides added to form the stem. B) Demonstration of visual detection using the molecular beacons. LAMP reactions with molecular beacons at a final concentration of 0.25 µM were incubated using a heat block and the endpoint fluorescence was visualized using a low cost viewer (miniPCR p51) and cell phone camera (Google Pixel 2 “Night Sight” mode).

We then tested the S6970 primers and beacons on synthetic RNA from wild-type (Twist Australia/VIC01/2020) and B.1.1.7 (Twist England/205041766/2020) SARS-CoV-2 variants. For comparison, we also multiplexed S6970 with the N2 primer set^11^ and molecular beacon ^12^. For these experiments, S6970 B117 MB was synthesized with a fluorescein (Fam) label, S6970 WT MB with a cyanine 3 (Cy3) label and N2 MB with a cyanine (Cy5) label.

Reactions were performed using locally produced enzyme mix equivalent to commercial RT-LAMP mix ^12^. Reactions consisted of 10 µL of 2x RT-LAMP enzyme mix, 2 µL of 10x primer-beacon mix and 8 µL of water doped with varying amounts of synthetic RNA. 10x primer-beacon mix consisted of 16 µM FIP, 16 µM BIP, 4 µM LF, 4 µM LB, 2 µM B3, 2 µM F3, 2.5 µM S6970 WT MB beacon and 2.5 µM S6970 B117 MB beacon. For multiplexed S6970 and N2 primer-beacon mix, the primer concentrations were halved and beacon N2 MB was added while maintaining all beacon concentrations at 2.5 µM.

Reactions were incubated for 60 minutes at 63^*°*^ C using a QuantStudio 5 qPCR machine for real-time quantification or a simple heat block. Note that the long incubation time was used to verify that no false positives were observed and most reactions completed within 30 minutes. We also used a simple and affordable (*∼*$25) illuminator (miniPCR p51) using blue light emitting diodes and an orange filter to observe the endpoint fluorescence of these assays.

## Results

Our goal was to design two molecular beacons that could be combined in a single reaction to distinguish SARS-CoV-2 variants containing the S1Δ69-70 deletion. To test our design, we used synthetic SARS-CoV-2 RNA containing either a prototypical genome sequence from early in the pandemic or a genome containing S1Δ69-70 deletion. We saw that fluorescence of two molecular beacons, S6970 WT MB and S6970 B117 MB, was easily visible by eye or cell phone camera after isothermal RT-LAMP and that the distinction between yellow Cy3 and green Fam allowed visual characterization of SARS-CoV-2 variants (Figure 1B).

To characterize the sensitivity and replicability of this assay, we attempted to amplify across a range of 10,000 to 39 genome copies per reaction (Figure 2A). The S6970 beacons were specific to their intended target with only minimal fluorescence detected from S6970 WT MB when Twist B.1.1.17 was amplified and only minimal fluorescence detected from S6970 B117 MB when wild type Twist was amplified. In contrast, in the presence of their intended targets, the beacons were strongly fluorescent. For comparison, the S6970 primer set was also multiplexed with the sensitive N2 primer set and molecular beacon. The N2 beacon was highly fluorescent for all positive samples (Figure 2B). In contrast, the S6970 primer set was less sensitive only detecting 11/12 reactions containing 625 genomic copies, 10/12 containing 312 copies, 9/12 containing 156 copies, 6/12 containing 78 copies and 5/12 containing 39 copies. No false positives were observed in any beacon.

**Figure 2.**
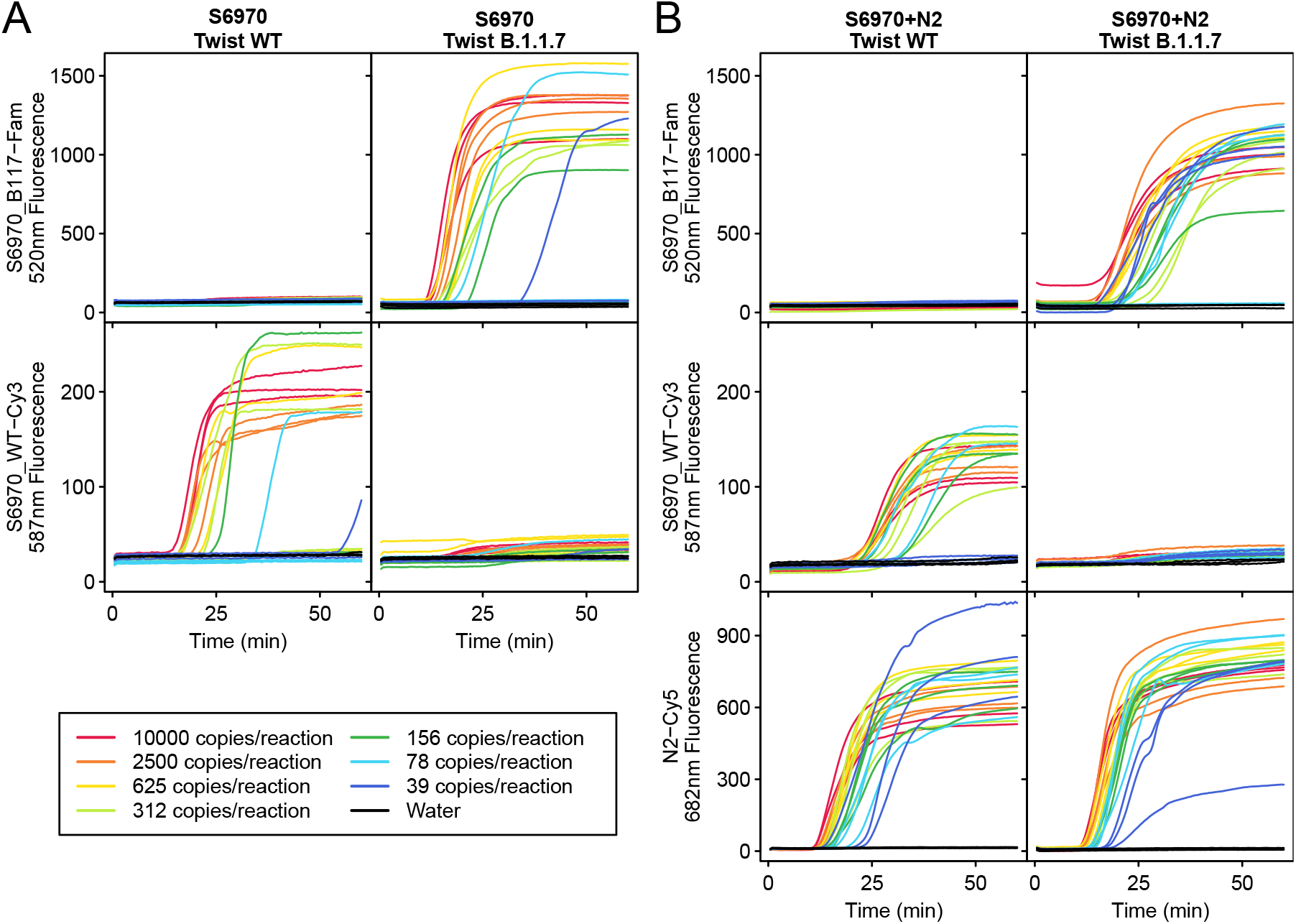
Testing of the S6970 primer set and molecular beacons on synthetic SARS-CoV-2 RNA. Fluorescence was monitored over time for LAMP isothermal amplification using the S6970 primer set alone or S6970 multiplexed with N2 primer for Twist Australia/VIC01/2020 (Twist WT) and Twist England/205041766/2020 (Twist B.1.1.7). The amount of synthetic SARS-CoV-2 RNA in each reaction was varied from 10,000 to 39 copies (indicated by line color) or water control (black). All fluorescence is given as relative fluorescence units (RFU) divided by 1000.

Estimating the probability of detecting a single copy of target RNA as *p*(positive| copies, detectionRate) = 1*—* (1*−* detectionRate)^copies^, the maximum likelihood estimate for the detection of a single copy for S6970 was 0.7% and the copy number necessary to give a 95% detection rate was estimated at 430 copies. With 8 µL of saliva input to each reaction, this translates to a detection at 54 copies per µL of saliva. This is less sensitive than the best SARS-CoV-2 primer sets, for example the N2 primer set used here, so would not be optimal for use as the only primer set in first line screening. But the primer set would be well suited to a follow up test of potential positives or multiplexed with more sensitive primer sets (Figure 2B). Due to the dual beacon design, the S6970 assay gives a clear readout of either WT SARS-CoV-2 detection or S1Δ69-70 detection or failure to detect either. Failures to detect SARS-CoV-2 in samples that are believed to be positive can be followed up with larger or more numerous reactions to give a higher probability of detection.

## Discussion

Here we show that molecular beacons and LAMP provide an effective method for detecting SARS-CoV-2 variants containing the S1Δ69-70 deletion. Using simple isothermal amplification of unpurified saliva and endpoint fluorescence, the genotype of a viral sample can be determined visually without expensive equipment. With locally-purified enzymes ^12^, the reagents costs can be reduced to pennies per test. Combined with the easily parallelizable nature of isothermal incubation, scaling is effectively limited only by sample collection and processing. Although the large deletion found in this variant was particularly well suited for detection, molecular beacons are often used to detect even single nucleotide polymorphisms ^13^. This pilot study showcases the capabilities of RT-LAMP and molecular beacons and promises further improvements in the monitoring and control of variants of concern through the design of additional beacons targeting diagnostic genetic signatures.

## Data Availability

All relevant data is included in the paper.

## Acknowledgements

This work was funded by the Penn Center for Research on Coronaviruses and Other Emerging Pathogens. We thank Young Hwang, Aoife Roche and members of the COVID-SAFE laboratory for help and suggestions.

## Notes

### Competing Interest Statement

The authors have declared no competing interest.

### Author Declarations

University of Pennsylvania Institutional Review Board

